# Safety and Feasibility of Serial Lumbar Punctures: Long-term Results from the Parkinson’s Progression Markers Initiative

**DOI:** 10.1101/2025.05.05.25327008

**Authors:** William Barbosa, Ruth B Schneider, Seung Ho Choi, Micah J Marshall, David-Erick Lafontant, Chelsea Caspell-Garcia, Christopher S Coffey, Jason Ross, Andrew Siderowf, Kenneth Marek, Tanya Simuni, The Parkinson’s Progression Markers Initiative (PPMI)

## Abstract

**Background and Objectives:** The collection of cerebrospinal fluid (CSF) serves an essential role in biomarker research. New Parkinson’s disease (PD) classifications include CSF α-synuclein status as a key biological anchor to enrich research trial design. Previous reports have established the safety of lumbar punctures (LPs) at baseline, but further investigation of longitudinal LP feasibility is needed. This study aimed to evaluate the safety and feasibility of serial CSF collection in participants enrolled in the Parkinson’s Progression Markers Initiative (PPMI).

**Methods:** PPMI participants were evaluated over a 13-year period. Descriptive statistics were calculated for all scheduled LPs occurring annually from baseline through year five and biennially thereafter. Adverse events were examined for all participants who attempted at least one LP. Compliance, defined as percentage of LPs with CSF collection, was assessed at baseline and for each longitudinal follow up visit. Logistic regression and generalized linear mixed effects models were used to calculate odds ratios and 95% confidence intervals for predictors of baseline and longitudinal LP success.

**Results:** 3479 participants enrolled in the PD (n=1412), prodromal (n=1768), and healthy control (n=299) cohorts were analyzed. 3360 participants attempted at least one LP of which 29.5% experienced an adverse event with 1.3% rated as severe. Compliance was 90% at baseline, 76.3% at year one, 67.8% at year three, 56.2% at year five, and 35.7% at year nine. From baseline to year five, percent change in compliance decreased by 39.4% in the PD cohort, 41.4% in the prodromal cohort, and 27.8% in the healthy control cohort. Predictive variables of baseline LP success included fewer years since diagnosis in the PD cohort (OR 0.82, 0.76-0.89), lower BMI in the prodromal cohort (OR 0.92, 0.89-0.94), and site location (U.S. vs. non-U.S.) for both PD (OR 1.5, 1.03-2.18) and healthy control (OR 3.6, 1.22-10.64) cohorts. Baseline LP success was the best predictor of longitudinal LP success (OR 7.82, 5.74-10.65).

**Discussion:** In this large study of longitudinal CSF collection, serial lumbar punctures were safe in PD research participants over a 13-year period. Although compliance was high over the first three years, further investigation is warranted to improve long term LP success.

## Introduction

The collection of cerebrospinal fluid (CSF) serves an essential role in the investigation of biomarkers of neurodegenerative disease[1–5]. Pathologic changes in the brain are reliably expressed in the CSF, often before clinical symptoms become manifest making this an ideal source for biological disease characterization[6–8]. Interventional studies are increasingly incorporating CSF biomarker metrics as part of their trial design[9–11] and it is anticipated that CSF collection will serve an important role in recruiting populations of interest in future disease prevention trials [12–14]. Historically Parkinson’s disease (PD) has been a clinical diagnosis, but the recent development of an α-synuclein seed amplification assay (SAA) marks a significant breakthrough[15–17].

Newly proposed biological definitions and research classifications of PD include CSF α-synuclein SAA status as a key biological anchor[18–20]. The collection of CSF via lumbar punctures (LPs) is expected to become integral to PD clinical trials and potentially in clinical practice as we continue to characterize and expand upon our understanding of biomarker assays[21, 22].

Currently, the CSF-SAA is being implemented to enrich research trial enrollment and identify participants who may benefit from targeted disease modifying intervention[12, 23–25]. Longitudinal CSF collection can be utilized to assess target engagement, measure therapeutic efficacy over time, and identify co-pathological biomarkers of disease progression[6, 26, 27].

As progress is made towards PD prevention trials, it remains imperative to understand the safety and feasibility of longitudinal LPs among pre-symptomatic individuals at high-risk for progression. While previous reports have established the safety and feasibility of LPs at baseline in the Parkinson’s Progression Markers Initiative (PPMI)[28], analysis of prodromal cohorts and longitudinal investigation is needed. Here we evaluate the safety and feasibility of serial CSF collection in participants with PD, without PD, and with prodromal disease status enrolled in PPMI. In addition, we examine demographic, clinical, and LP-related characteristics to identify potential predictors of longitudinal LP success to help inform future clinical trial design.

## Methods

PPMI is an ongoing observational, multi-center, international study designed to identify clinical, genetic, imaging, and biological markers of PD progression to support advances in treatment. The PPMI study design, methods, and eligibility criteria have been reported previously[29] and are available at www.ppmi-info.org/study-design. Participants undergo LPs annually from baseline through year five and biennially thereafter with CSF collection performed predominantly using 24-gauge needles by trained study site investigators.

For the purposes of this study, we analyzed data from PPMI participants enrolled in the PD (sporadic, leucine rich repeat kinase 2 (LRRK2), glucocerebrosidase (GBA), rare genetic), prodromal (REM sleep behavior disorder (RBD), hyposmia, LRRK2, GBA, rare genetic), and healthy control cohorts. LPs were conducted over a 13-year period across all scheduled LP visits. Baseline demographic variables were summarized and stratified by subgroup to include age, sex, BMI, race, and ethnicity. Clinical variables included the number of years since diagnosis, Movement Disorders Society Unified Parkinson’s Disease Rating Scale I-III scores, Hoehn & Yahr, and Montreal Cognitive Assessment. CSF collection metrics examined included total fluid volume, incidence of traumatic LPs, and fluoroscopy utilization.

Adverse events (AEs) related to LPs were examined for all participants who attempted at least one LP procedure. The occurrence of AEs was assessed at the time of the study visit and again within one week following the procedure. AEs were summarized by event type, site-reported severity, and cohort. Compliance, defined as CSF collection status of “Collected” or “Partial Collection” in eligible participants, was examined at baseline and upon longitudinal follow up. Reasons for CSF collection not completed were further evaluated by cohort and study visit.

To assess the association between baseline and longitudinal success, odds ratios, associated 95% Wald confidence intervals and p-values comparing the odds of at least one compliant post-baseline visit given baseline compliance vs. non-compliance were computed. To identify specific baseline predictors of LP success, two sets of models were run for each cohort: logistic regression models to predict baseline success, and generalized linear mixed effects models with binomial distribution and logit link to predict longitudinal LP success. Candidate predictors included age, sex, years since diagnosis (PD cohort only), BMI, race, ethnicity, site (US vs. non-US), and a two-way interaction between sex and BMI. Interactions with time were also considered in the longitudinal models. Utilizing an initial screening step that included covariates that were significant at α = 0.10 in univariate/screening models, a backward selection procedure was employed to sequentially remove variables until all remaining variables (or their interaction with time) were significant at α = 0.05. Longitudinal models with LP method (fluoroscopy vs. non-fluoroscopy) and occurrence of LP-related AE within 7 days of baseline LP as predictors were run separately in the subset of participants who had a successful baseline LP. Odds ratios, associated 95% confidence intervals, and p-values are reported in the final models. Longitudinal models were estimated using Laplace estimation with unstructured covariance structure and inclusion of random intercept and slope terms. Statistical analyses were performed using SAS v9.4 (SAS Institute Inc., Cary, NC, USA; sas.com; RRID:SCR_008567). Data was downloaded on June 17, 2024.

## Results

3479 PPMI participants enrolled in the PD (n=1412), prodromal (n=1768), and healthy control (n=299) cohorts were analyzed. Baseline age, sex, race, and BMI variables were comparable across cohorts (median age of 65 years, 55% male, 94% White, and median BMI 26.6). For all study participants baseline LP compliance with either collected or partially collected CSF was 90%, of which 13% were fluoroscopy guided and 10% were traumatic. Lower baseline compliance percentages of 78% and 66% were observed in the GBA PD and rare genetic variant PD subgroups respectively. Further baseline demographics and clinical characteristics of study participants stratified by subgroup are summarized in **Table 1**.

**Table 1.**
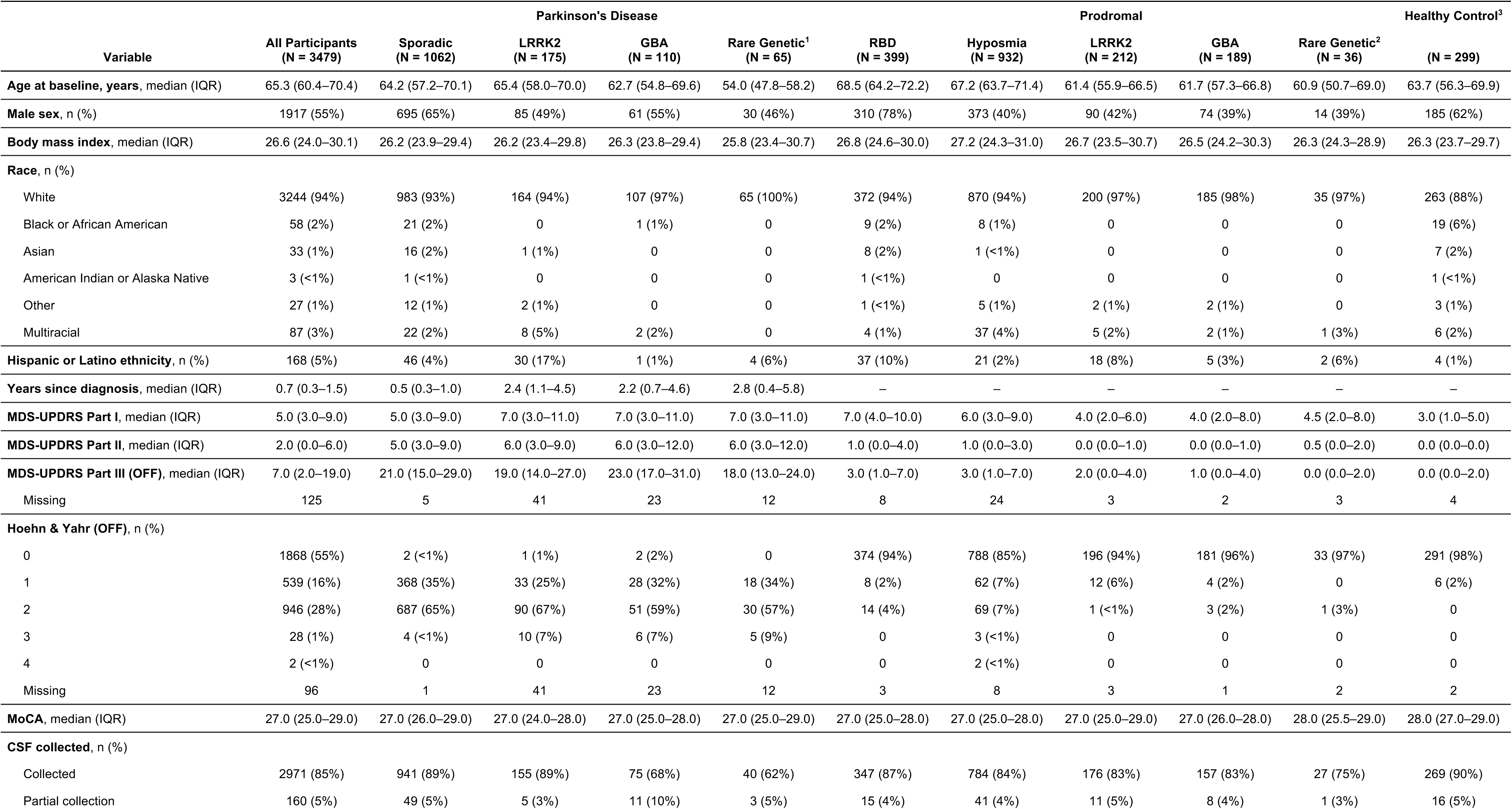
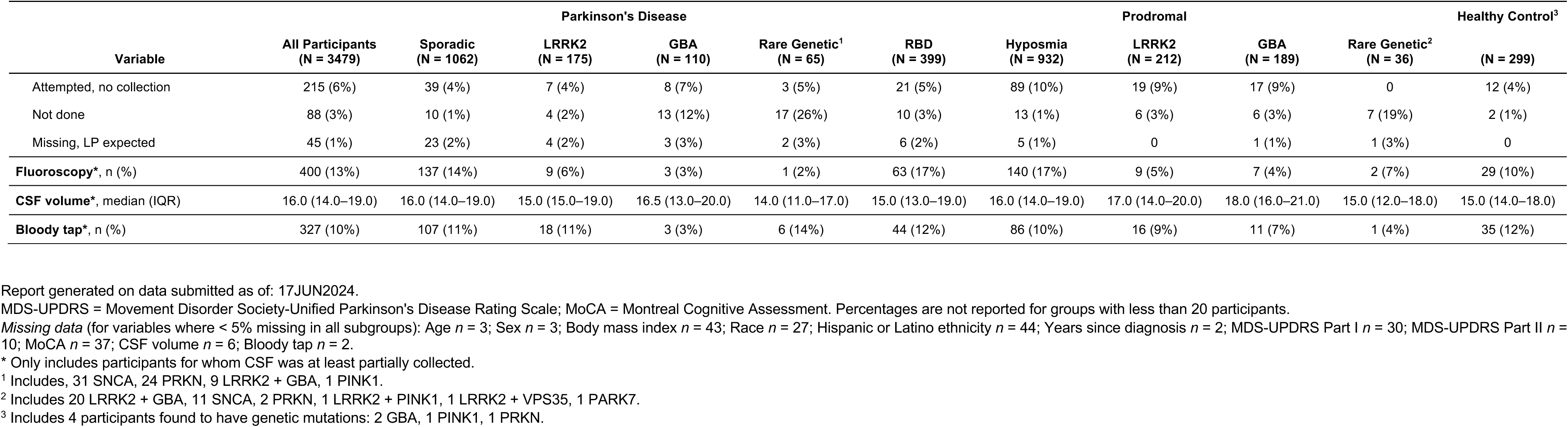
Baseline demographics, disease characteristics, and lumbar puncture status.

Among all participants who attempted at least one LP (n=3360), 29.5% experienced an adverse event (**Table 2**). The most common adverse events were headache (16.0%) and back/injection site pain (14.1%). The healthy control cohort exhibited a higher proportion of adverse events (44.6%) when compared to PD (31.4%) and prodromal (25.3%) cohorts. Examination of adverse event severity of all participants revealed that 22.9% (n=769) experienced a mild event, 8.9% (n=299) experienced a moderate event, and 1.3% (n=43) experienced a severe event.

**Table 2.** Lumbar puncture-related adverse events by type, severity and cohort.

| Adverse Event | All Participants (N = 3360) |  |  |  | Parkinson's Disease (N = 1341) |  |  |  | Prodromal (N = 1721) |  |  |  | Healthy Control (N = 298) |  |  |  |
| --- | --- | --- | --- | --- | --- | --- | --- | --- | --- | --- | --- | --- | --- | --- | --- | --- |
|  | # of Ps. | % of Ps. | # of Events | Rate | # of Ps. | % of Ps. | # of Events | Rate | # of Ps. | % of Ps. | # of Events | Rate | # of Ps. | % of Ps. | # of Events | Rate |
| Total | 990 | 29.5% | 1669 | 0.021 | 421 | 31.4% | 707 | 0.018 | 436 | 25.3% | 719 | 0.027 | 133 | 44.6% | 243 | 0.017 |
| Mild | 769 | 22.9% | 1234 | 0.015 | 323 | 24.1% | 500 | 0.013 | 350 | 20.3% | 571 | 0.021 | 96 | 32.2% | 163 | 0.012 |
| Moderate | 299 | 8.9% | 384 | 0.005 | 134 | 10.0% | 182 | 0.005 | 116 | 6.7% | 136 | 0.005 | 49 | 16.4% | 66 | 0.005 |
| Severe | 43 | 1.3% | 51 | 0.0006 | 22 | 1.6% | 25 | 0.0006 | 11 | 0.6% | 12 | 0.0004 | 10 | 3.4% | 14 | 0.001 |
| Headache | 538 | 16.0% | 666 | 0.008 | 242 | 18.0% | 321 | 0.008 | 226 | 13.1% | 255 | 0.009 | 70 | 23.5% | 90 | 0.006 |
| Mild | 367 | 10.9% | 444 | 0.006 | 167 | 12.5% | 207 | 0.005 | 159 | 9.2% | 181 | 0.007 | 41 | 13.8% | 56 | 0.004 |
| Moderate | 175 | 5.2% | 190 | 0.002 | 84 | 6.3% | 98 | 0.002 | 64 | 3.7% | 65 | 0.002 | 27 | 9.1% | 27 | 0.002 |
| Severe | 31 | 0.9% | 32 | 0.0004 | 15 | 1.1% | 16 | 0.0004 | 9 | 0.5% | 9 | 0.0003 | 7 | 2.3% | 7 | 0.0005 |
| Back or injection site pain | 475 | 14.1% | 667 | 0.008 | 176 | 13.1% | 236 | 0.006 | 230 | 13.4% | 319 | 0.012 | 69 | 23.2% | 112 | 0.008 |
| Mild | 411 | 12.2% | 554 | 0.007 | 153 | 11.4% | 196 | 0.005 | 200 | 11.6% | 278 | 0.010 | 58 | 19.5% | 80 | 0.006 |
| Moderate | 89 | 2.6% | 106 | 0.001 | 32 | 2.4% | 38 | 0.0010 | 37 | 2.1% | 40 | 0.001 | 20 | 6.7% | 28 | 0.002 |
| Severe | 7 | 0.2% | 7 | 0.0001 | 2 | 0.1% | 2 | 0.0001 | 1 | 0.1% | 1 | <.0001 | 4 | 1.3% | 4 | 0.0003 |
| Other | 260 | 7.7% | 336 | 0.004 | 114 | 8.5% | 150 | 0.004 | 111 | 6.4% | 145 | 0.005 | 35 | 11.7% | 41 | 0.003 |
| Mild | 196 | 5.8% | 236 | 0.003 | 80 | 6.0% | 97 | 0.002 | 92 | 5.3% | 112 | 0.004 | 24 | 8.1% | 27 | 0.002 |
| Moderate | 69 | 2.1% | 88 | 0.001 | 35 | 2.6% | 46 | 0.001 | 24 | 1.4% | 31 | 0.001 | 10 | 3.4% | 11 | 0.0008 |
| Severe | 10 | 0.3% | 12 | 0.0001 | 6 | 0.4% | 7 | 0.0002 | 2 | 0.1% | 2 | 0.0001 | 2 | 0.7% | 3 | 0.0002 |

Report generated on data submitted as of: 17JUN2024.
Column definitions:

Longitudinal examination across cohorts revealed compliance was 90% at baseline, 76.3% at year one, 67.8% at year three, 56.2% at year five, and 35.7% at year nine (**Table 3**). From baseline to year five, percent change in compliance decreased by 39.4% in the PD cohort, 41.4% in the prodromal cohort, and 27.8% in the healthy control cohort. The greatest drop off in compliance rates occurred between the baseline visit and year one, and between years five and seven (**Figure 1**).

**Figure 1:**
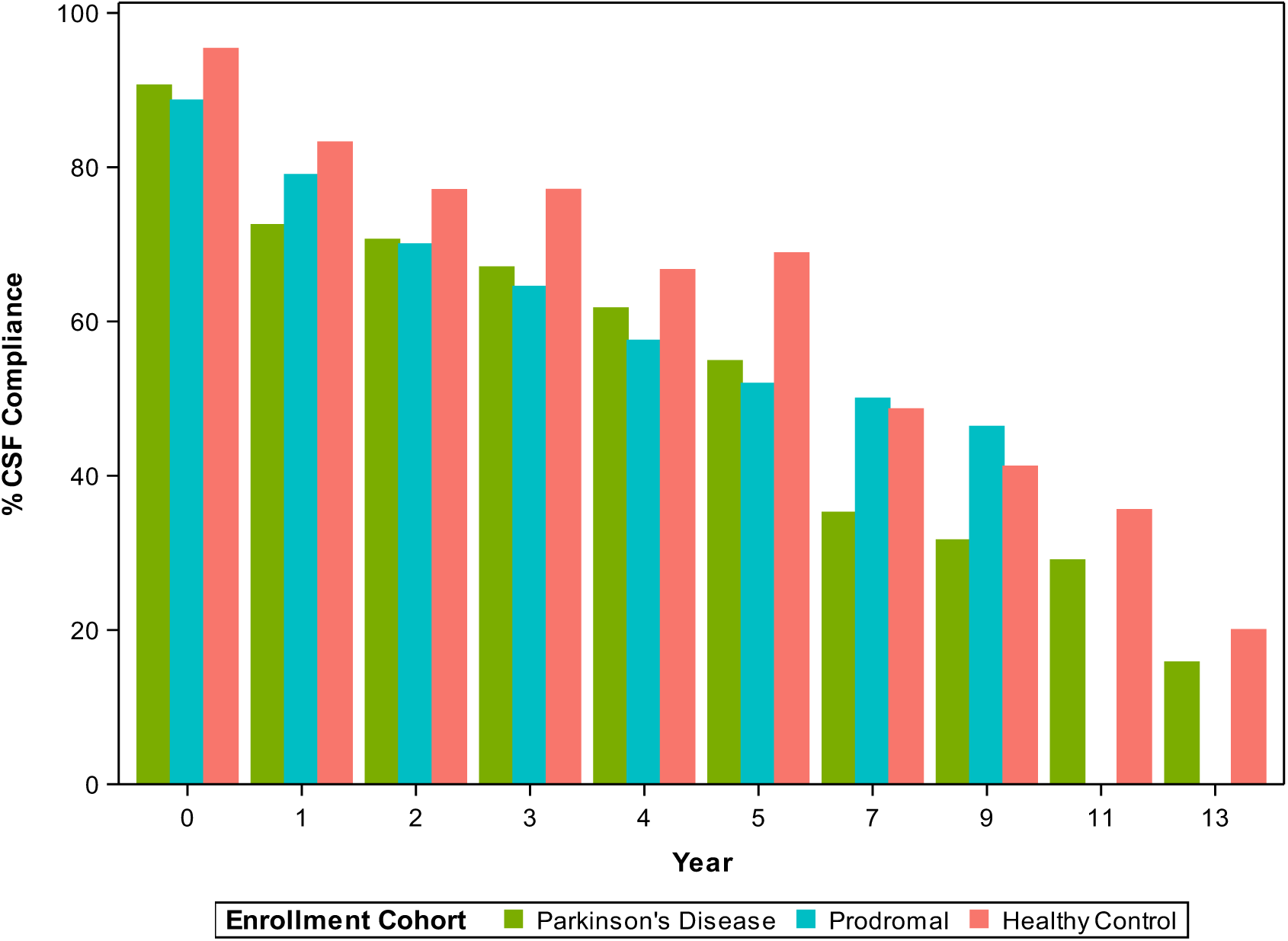
Longitudinal CSF Compliance by Cohort

**Table 3.**
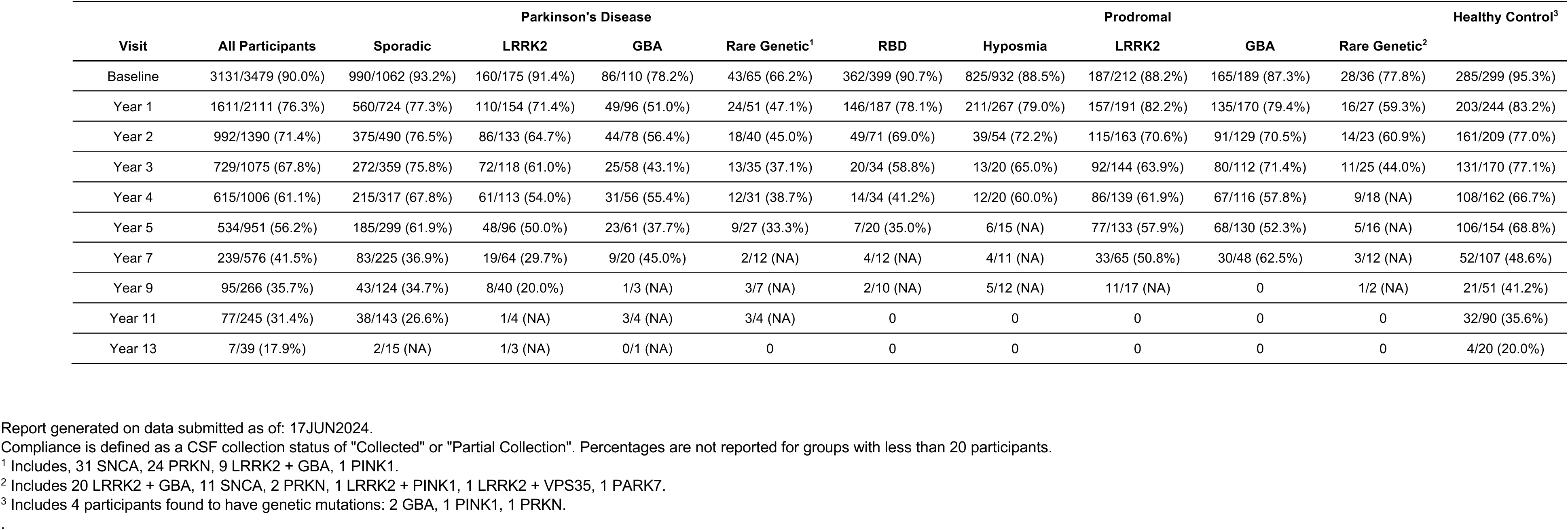
Longitudinal CSF compliance by group.

Odds ratios and 95% confidence intervals for covariant predictors of baseline LP success were calculated with statistically significant results shown in **Table 4**. Baseline variables of significance included years since diagnosis in the PD cohort (OR 0.82, 0.76-0.89), BMI in the prodromal cohort (OR 0.92, 0.89-0.94), and site location (U.S. vs. non-U.S.) for both PD (OR 1.50, 1.03-2.18) and healthy control (OR 3.60, 1.22-10.64) cohorts.

**Table 4.**
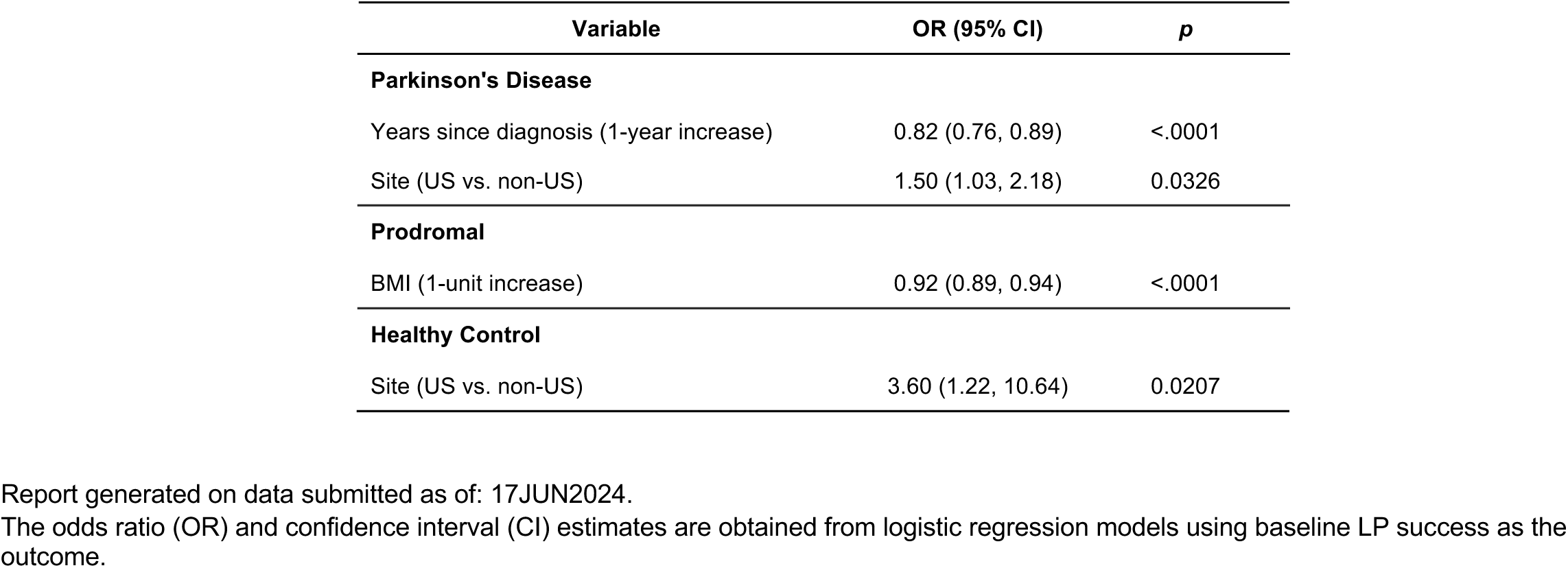
Predictors of baseline lumbar puncture success.

Baseline LP success was the most significant predictor of longitudinal LP success (OR 7.82, 5.74-10.65) (**Supplemental Table 1**). Across all cohorts, time from baseline predicted lower likelihood for longitudinal LP success with odds ratios of 0.46 (0.43-0.50), 0.66 (0.58-0.76), and 0.51 (0.45-0.59) in PD, prodromal, and healthy control cohorts respectively (**Table 5**). For participants with a successful baseline LP, the occurrence of an LP-related AE within 7 days of baseline predicted lower likelihood of longitudinal LP success in the PD (OR 0.25, 0.11 – 0.56) and prodromal (OR 0.28, 0.15-0.52) cohorts. Amongst those who did not undergo a successful LP, participant refusal was the most cited reason for which CSF was not collected across all cohorts (**Supplemental Table 2**).

**Table 5.** Predictors of longitudinal lumbar puncture success.

| Variable | OR (95% CI) | p |
| --- | --- | --- |
| **Parkinson's Disease** |  |  |
| Time (1-year increase) | 0.46 (0.43, 0.50) | <.0001 |
| Years since diagnosis (1-year increase) | 0.74 (0.66, 0.82) | <.0001 |
| Site (US vs. non-US) | 3.15 (2.08, 4.77) | <.0001 |
| **For participants with successful baseline LP only (separate model)** |  |  |
| Occurrence of LP-related AE within 7 days of baseline LP | 0.25 (0.11, 0.56) | 0.0008 |
| Baseline LP-related AE x time interaction | 1.29 (1.10, 1.51) | 0.0018 |
| **Prodromal** |  |  |
| Time (1-year increase)* | 0.66 (0.58, 0.76) | <.0001 |
| Difference for a 1-year increase in age | 0.99 (0.97, 1.00) | 0.0397 |
| Difference for US vs. non-US site | 1.45 (1.12, 1.88) | 0.0044 |
| Age (1-year increase) | 1.00 (0.98, 1.03) | 0.7275 |
| BMI (1-unit increase) | 0.94 (0.91, 0.97) | 0.0003 |
| Site (US vs. non-US) | 1.61 (1.02, 2.54) | 0.0410 |
| **For participants with successful baseline LP only (separate models)** |  |  |
| Baseline LP method (fluoroscopy vs. non-fluoroscopy) | 0.38 (0.16, 0.93) | 0.0338 |
| Occurrence of LP-related AE within 7 days of baseline LP | 0.28 (0.15, 0.52) | <.0001 |
| **Healthy Control** |  |  |
| Time (1-year increase) | 0.51 (0.45, 0.59) | <.0001 |
| Sex (male vs. female) | 2.42 (1.07, 5.47) | 0.0331 |
| Site (US vs. non-US) | 3.42 (1.32, 8.85) | 0.0112 |
| **For participants with successful baseline LP only (separate model)** |  |  |
| Baseline LP method (fluoroscopy vs. non-fluoroscopy) | 0.02 (0.002, 0.19) | 0.0006 |

Report generated on data submitted as of: 17JUN2024.
The odds ratio (OR) and confidence interval (CI) estimates are obtained from generalized linear mixed models using LP success at each annual visit as the outcome.
\*Due to the inclusion of interaction effects, the time estimate is calculated for a participant of average age (64.9 years) based at a US site.

## Discussion

In this large study of longitudinal CSF collection, serial lumbar punctures were safe across PD, prodromal PD, and healthy control research participants over a 13-year period of observation. Most adverse events were of mild severity and only approximately 1% of events were rated as severe. The most common adverse events were headache and back/site injection pain, which is in accordance with other multi-center baseline LP studies[30, 31].

Baseline compliance for all participants was high with lower rates appreciated in the genetic cohort subgroups. This is likely due to their less stringent requirements for enrollment in PPMI as genetic cohort participants were allowed to be recruited despite LP refusal in attempt to maximize feasibility of study recruitment. Favorable predictors of baseline LP success were shorter interval time from diagnosis, having a lower BMI, and study site location in the United States. While prior studies have established a correlation between higher BMI and unsuccessful LP outcomes[32, 33], the effects of time from diagnosis and study site location are not yet fully understood. Presumably greater time from diagnosis may result in additional comorbidities that could influence LP success. Additionally, there may be unrecognized study site differences in LP technique or informed consent procedures that could also impact success rates, but this requires further examination.

Over the first three years, longitudinal compliance was high but subsequently exhibited steady attrition across cohorts over time. Participant refusal remained the most cited reason for unsuccessful CSF collection, but other potential contributors to consider include loss of motivation, escalating symptom burden, lack of time/resources, or prior negative LP experience. Interestingly, healthy controls demonstrated the highest baseline and longitudinal compliance despite having a greater percentage of adverse events. Amongst PD and prodromal participants who underwent a successful baseline LP, those who experienced any adverse event within 7 days of baseline CSF collection demonstrated lower longitudinal compliance. These results further highlight the importance of maximizing baseline LP success as this was the best predictor of long-term compliance. To ensure high compliance rates PPMI committed substantial resources to training sites and standardization of procedures. Additional implemented strategies included providing standardized information packages and videos to participants on the process of CSF fluid collection, and establishing a centralized LP education core to offer consistent information at the participant friendly level.

While notable strengths of this study include the large sample size, extended duration of follow up, and comprehensive evaluation of factors associated with successful LPs, there are also limitations worth acknowledging. First there was lack of racial and ethnic diversity among the enrolled participants. Additionally, while rating of adverse event severity was graded according to commonly accepted research standards, some subjectivity in assessment and report of symptoms remains. Similarly, the determination as to which participants were selected to undergo fluoroscopy occurred at the site level and was not standardized. Examination of site level differences was restricted to whether or not CSF collection took place in the United States. Although participant refusal was the most cited reason for unsuccessful CSF collection, data were not collected to understand the reason for refusal providing limited opportunity to understand the rationale for declining the procedure. This limitation is compounded by the significant percentage of responses for which no reason was provided for why CSF was not collected. Lastly, within the prodromal and genetic subgroup cohorts there is shorter duration of follow up as recruitment of these groups was initiated a few years following study launch.

In conclusion, although the safety profile for LPs in PPMI is reassuring and compliance for serial LPs is high over the first three years, further investigation is warranted to explore methods to improve long-term retention. Efforts to better understand the rationale for participant refusal and examination of site level differences may highlight how best to minimize study attrition moving forward. Clarifying identified participant uncertainties or misconceptions, readdressing best practices for the process of informed consent, and establishing more comprehensive site training on procedural technique may prove to be effective methods to ensure longitudinal LP success. While it remains imperative to examine methods to improve long term participant retention, this study provides promising insights to help optimize serial CSF collection and inform clinical trial design moving forward.

## Data Availability

Data used in the preparation of this article were obtained on June 17, 2024 from the Parkinson’s Progression Markers Initiative (PPMI) database (www.ppmi-info.org/access-data-specimens/download-data), RRID:SCR_006431. For up-to-date information on the study, visit www.ppmi-info.org. This analysis was conducted by the PPMI Statistics Core and used actual dates of activity for participants, a restricted data element not available to public users of PPMI data. Protocol information for The Parkinson’s Progression Markers Initiative (PPMI) Clinical - Establishing a Deeply Phenotyped PD Cohort AM 3.2. can be found on protocols.io or by following this link: https://dx.doi.org/10.17504/protocols.io.n92ldmw6ol5b/v2. Statistical analysis codes used to perform the analyses in this article are shared on Zenodo (10.5281/zenodo.15079775).

## Acknowledgements

This study was funded through support from the Michael J. Fox Foundation for Parkinson’s Research Write Now Initiative. PPMI – a public-private partnership – is funded by the Michael J. Fox Foundation for Parkinson’s Research and funding partners, including 4D Pharma, Abbvie, AcureX, Allergan, Amathus Therapeutics, Aligning Science Across Parkinson’s, AskBio, Avid Radiopharmaceuticals, BIAL, BioArctic, Biogen, Biohaven, BioLegend, BlueRock Therapeutics, Bristol-Myers Squibb, Calico Labs, Capsida Biotherapeutics, Celgene, Cerevel Therapeutics, Coave Therapeutics, DaCapo Brainscience, Denali, Edmond J. Safra Foundation, Eli Lilly, Gain Therapeutics, GE HealthCare, Genentech, GSK, Golub Capital, Handl Therapeutics, Insitro, Jazz Pharmaceuticals, Johnson & Johnson Innovative Medicine, Lundbeck, Merck, Meso Scale Discovery, Mission Therapeutics, Neurocrine Biosciences, Neuron23, Neuropore, Pfizer, Piramal, Prevail Therapeutics, Roche, Sanofi, Servier, Sun Pharma Advanced Research Company, Takeda, Teva, UCB, Vanqua Bio, Verily, Voyager Therapeutics, the Weston Family Foundation and Yumanity Therapeutics.

## Conflicts of Interest and Disclosures

RBS declares research funding from the National Institutes of Health, Michael J. Fox Foundation for Parkinson’s Research, Parkinson’s Foundation, Bial, Biohaven Pharmaceuticals, and Acadia Pharmaceuticals. She has participated on a Sun Pharma Advisory Board and received funding from Clintrex Research Corporation for participation on a DSMB SHC declares research funding to his institution from The Michael J. Fox Foundation and NIH/NINDS MM declares research funding to his institution from The Michael J. Fox Foundation DEL declares research funding to his institution from The Michel J. Fox Foundation CCG declares research funding to her institution from The Michael J. Fox Foundation CSC declares grants from The Michael J. Fox Foundation and NIH/NINDS

## Notes

### Competing Interest Statement

RBS declares research funding from the National Institutes of Health, Michael J. Fox Foundation for Parkinsons Research, Parkinsons Foundation, Bial, Biohaven Pharmaceuticals, and Acadia Pharmaceuticals. She has participated on a Sun Pharma Advisory Board and received funding from Clintrex Research Corporation for participation on a DSMB SHC declares research funding to his institution from The Michael J. Fox Foundation and NIH/NINDS MM declares research funding to his institution from The Michael J. Fox Foundation DEL declares research funding to his institution from The Michel J. Fox Foundation CCG declares research funding to her institution from The Michael J. Fox Foundation CSC declares grants from The Michael J. Fox Foundation and NIH/NINDS

### Clinical Protocols

https://www.ppmi-info.org/study-design

### Funding Statement

This study was funded through support from the Michael J. Fox Foundation for Parkinsons Research Write Now Initiative.
PPMI a public-private partnership is funded by the Michael J. Fox Foundation for Parkinsons Research and funding partners, including 4D Pharma, Abbvie, AcureX, Allergan, Amathus Therapeutics, Aligning Science Across Parkinsons, AskBio, Avid Radiopharmaceuticals, BIAL, BioArctic, Biogen, Biohaven, BioLegend, BlueRock Therapeutics, Bristol-Myers Squibb, Calico Labs, Capsida Biotherapeutics, Celgene, Cerevel Therapeutics, Coave Therapeutics, DaCapo Brainscience, Denali, Edmond J. Safra Foundation, Eli Lilly, Gain Therapeutics, GE HealthCare, Genentech, GSK, Golub Capital, Handl Therapeutics, Insitro, Jazz Pharmaceuticals, Johnson & Johnson Innovative Medicine, Lundbeck, Merck, Meso Scale Discovery, Mission Therapeutics, Neurocrine Biosciences, Neuron23, Neuropore, Pfizer, Piramal, Prevail Therapeutics, Roche, Sanofi, Servier, Sun Pharma Advanced Research Company, Takeda, Teva, UCB, Vanqua Bio, Verily, Voyager Therapeutics, the Weston Family Foundation and Yumanity Therapeutics.

### Author Declarations

Data used in the preparation of this article were obtained on June 17, 2024 from the Parkinsons Progression Markers Initiative (PPMI) database (www.ppmi-info.org/access-data-specimens/download-data), RRID:SCR_006431.

